# Vaporized Cannabis versus Placebo for Acute Migraine: A Randomized Controlled Trial

**DOI:** 10.1101/2024.02.16.24302843

**Authors:** Nathaniel M. Schuster, Mark S. Wallace, Thomas D. Marcotte, Dawn C. Buse, Euyhyun Lee, Lin Liu, Michelle Sexton

**Affiliations:** Center for Pain Medicine, Department of Anesthesiology, San Diego Health System, San Diego, CA; Department of Psychiatry, San Diego Health System, San Diego, CA; Altman Clinical and Translational Research Institute, San Diego Health System, San Diego, CA; Department of Biostatistics and Bioinformatics, Herbert Wertheim School of Public Health and Human Longevity Science, San Diego Health System, San Diego, CA; Centers for Integrative Health, Department of Family Medicine, San Diego Health System, San Diego, CA; Center for Medicinal Cannabis Research, University of California, San Diego Health System, San Diego, CA; Department of Neurology, Albert Einstein College of Medicine

**Author notes:** Corresponding author: Nathaniel M. Schuster, MD, UC San Diego Center for Pain Medicine, 9400 Campus Point Drive, Suite 1-300, La Jolla, CA 92037.

## Abstract

**Background:** Preclinical and retrospective studies suggest cannabinoids may be effective in migraine treatment. However, there have been no randomized clinical trials examining the efficacy of cannabinoids for acute migraine.

**Methods:** In this randomized, double-blind, placebo-controlled, crossover trial, adults with migraine treated up to 4 separate migraine attacks, 1 each with vaporized 1) 6% Δ9-tetrahydrocannabinol (THC-dominant); 2) 11% cannabidiol (CBD-dominant); 3) 6% THC+11% CBD; and 4) placebo cannabis flower in a randomized order. Washout period between treated attack was ≥1 week. The primary endpoint was pain relief and secondary endpoints were pain freedom and most bothersome symptom (MBS) freedom, all assessed at 2 hours post-vaporization.

**Results:** Ninety-two participants were enrolled and randomized, and 247 migraine attacks were treated. THC+CBD was superior to placebo at achieving pain relief (67.2% vs 46.6%, Odds Ratio [95% Confidence Interval] 2.85 [1.22, 6.65], p=0.016), pain freedom (34.5% vs. 15.5%, 3.30 [1.24, 8.80], p=0.017) and MBS freedom (60.3% vs. 34.5%, 3.32 [1.45, 7.64], p=0.005) at 2 hours, as well as sustained pain freedom at 24 hours and sustained MBS freedom at 24 and 48 hours. THC-dominant was superior to placebo for pain relief (68.9% vs. 46.6%, 3.14 [1.35, 7.30], p=0.008) but not pain freedom or MBS freedom at 2 hours. CBD-dominant was not superior to placebo for pain relief, pain freedom or MBS freedom at 2 hours. There were no serious adverse events.

**Conclusions:** Acute migraine treatment with 6% THC+11% CBD was superior to placebo at 2 hours post-treatment with sustained benefits at 24 and 48 hours.

**Trial Registration Number:** NCT04360044

**Disclosures:** NMS has received research funding from Migraine Research Foundation, Novaremed, UCSD Academic Senate, UCSD Department of Anesthesiology RAG, and NIH CTSA Grant UL1TR000100 and has consulted for Eli Lilly & Co., Averitas, Syneos, Schedule 1 Therapeutics, Vectura Fertin, and ShiraTronics. He serves on the editorial board of Pain Medicine (Oxford University Press). DCB has been a consultant for Abbvie, Amgen, Biohaven, Lilly, Lundbeck, Teva, Theranica. She has received grant support from the US Food and Drug Association and National Headache Foundation. She is an editor for Current Pain and Headache Reports. MSW, TM, EL, LL and MS have no disclosures to report.

## Introduction

Migraine is the second leading cause of years lived with disability worldwide. It affects over a billion people worldwide, including 38 million Americans^1^. Migraine treatments are classified for acute and/or preventive use^2^. Nearly everyone with migraine uses acute treatments for migraine attacks. However, rates of treatment optimization with traditional acute therapies are relatively low and rates of discontinuation are high^3,4^. There is significant interest in and use of cannabinoids for acute migraine treatment. Migraine is among the most common medicinal uses of cannabinoids, with 35.5% of 1,429 medical cannabis users reporting use for headache/migraine; inhalation was the most common route of administration (81.4%)^5^.

More than 125 identified phytocannabinoids are naturally found in the cannabis plant, including Δ9-tetrahydrocannabinol (THC) and cannabidiol (CBD). Although the US Food and Drug Administration (FDA) has approved cannabinoid-based medications for specific conditions, none have been approved for migraine. At the time of this writing, THC was legal for medical and/or recreational use in 38 US states and the District of Columbia. Since 2018, hemp-derived CBD (<0.3% THC) is not a controlled substance in the US^6^.

Patients often ask healthcare professionals about cannabinoids for migraine, but data to inform medical advice is lacking^7^. Preclinical evidence suggests that cannabinoids may have effects on migraine pathogenesis through mechanisms including inhibiting calcitonin gene-related peptide (CGRP) release, inhibiting CGRP-induced nitric oxide release, inhibiting trigeminovascular neurons, and inhibiting cortical spreading depression, and surveys and retrospective studies suggest that cannabinoids may have benefit as acute and/or preventive treatments for migraine^8–20^. However the efficacy of cannabis for acute treatment of migraine has not been studied via a randomized, controlled trial (RCT)^10,12,21^. Therefore, this RCT was conducted to assess the efficacy of vaporized cannabis against placebo cannabis.

## Methods

### Trial Design

This randomized, double-blind, placebo-controlled, crossover trial was conducted at the University of California, San Diego (UCSD). The study was approved by the UCSD Institutional Review Board (IRB #181944) and the US FDA. The study was prospectively registered, and full protocol is available, at ClinicalTrials.gov (NCT04360044). The authors take full responsibility for the data and the accuracy and integrity of the publication. No writing assistance was provided by outside parties. The study was performed in accordance with the principles of the Declaration of Helsinki. All participants provided written informed consent prior to enrollment.

### Sample Size

Sample size was determined based on our primary outcome of pain relief at 2 hours, assuming a response of 68% for treatment and 45% for placebo as seen in an intranasal sumatriptan RCT^22^. This effect size required 72 participants to detect a significant difference with 80%. Assuming a 20% dropout rate, we planned to enroll 90 participants.

### Study Drug

Cannabis flower was obtained from the National Institute on Drug Abuse (NIDA) Drug Supply Program (DSP) and consisted of 4 different treatments. The most closely matched batches available from the NIDA DSP at study initiation were selected. They were: 1) 5.62% THC+0.03% CBD (referred to in this study as 6% THC or THC-dominant); 2) 11.27% CBD+0.35% CBD (referred to as 11% CBD or CBD-dominant); 3) 6.16% THC + 10.77% CBD (referred to as 6% THC+11% CBD or THC+CBD); and 4) placebo flower (<0.025% THC + 0.14% CBD). All treatments contained <1% minor cannabinoids and were devoid of terpenes. The studied THC potency was based on previous pain studies^23–25^.

### Participants

Key inclusion criteria were: ages 21-65; migraine according to International Classification of Headache Disorders, 3^rd^ edition (ICHD-3) criteria^26^; 2-23 headache days and 2-23 migraine days per month; agree not to use cannabis (outside of the study drug), opioids or barbiturates. Key exclusion criteria were: screening visit urine drug test positive for THC, barbiturates, opioids, oxycodone, or methadone; pregnant; breastfeeding; known cognitive impairment; current moderate-severe or severe depression; history of bipolar disorder, schizophrenia, or psychosis; history of substance use disorder; and active pulmonary disease or other severe medical illnesses at the discretion of the researchers. Inclusion criteria initially required THC use within the prior 2 years; this was removed March 18, 2021 due to slow recruitment and no serious adverse events at that time. Full criteria are available at ClinicalTrials.gov.

Participants agreed not to use any other acute migraine treatments prior to or within 2 hours following study drug administration.

### Enrollment

Participants were enrolled at UCSD November 20, 2020-November 4, 2022 with follow-up completed February 23, 2023. A headache neurologist experienced in ICHD-3 criteria (NMS) confirmed participant eligibility. After written informed consent, baseline characteristics were captured using REDCap^27^. Then participants were trained in the Foltin Uniform Puff Procedure (FUPP), a validated cannabis vaporization procedure, and a smartphone application was installed on participants’ smartphones to guide participants through the study and collect data^28^.

### Randomization

A research pharmacist randomized participants to receive the 4 different treatments using simple 1:1:1:1 assignment (24 possible orders) using Microsoft Excel’s random number function.

### Blinding

Participants, research coordinators, investigators, and statisticians were blinded until after the initial statistical analysis was completed; only the research pharmacists were unblinded during the study. The four treatments were prepared into identical Storz & Bickel Filling Set vaporization capsules by a research pharmacist and placed into sealed bags. Labels were affixed to the sealed bags stating the order in which the participant would use the four treatments (“Migraine 1” through “Migraine 4”). The key linking the treatments to their identifying number was stored on a password-protected computer in the locked pharmacy available only to the research pharmacists.

Participant blinding was promoted by using placebo cannabis flower from which the THC and CBD had been extracted via a chemical process by the NIDA DSP and by framing, including educating patients that they might experience a “placebo high” from the CBD-dominant and placebo treatments and that they might not experience a “high” from the THC-dominant and THC+CBD treatments as the study potencies were lower than “recreational” potencies. Each attack, after answering 2-hour efficacy and safety questions, participants were asked which treatment they thought they received.

### Treatments

Upon each migraine attack onset, the participant accessed the smartphone application. If ≥7 days had elapsed since the prior cannabis administration (ensuring ≥7 days washout period between cannabis administrations) the application asked the participant questions to establish whether the attack met treatment criteria of: 1) headache <4 hours from onset, 2) headache pain moderate or severe in intensity, 3) associated with photophobia and phonophobia and/or with nausea, 4) no acute treatments used since attack onset. If the attack met all criteria, the application instructed the participant to vaporize study cannabis at 180° Celsius and guided the participant through the timed FUPP to standardize vaporization procedure across treated attacks and across participants. FUPP consists of five seconds of inhalation, followed by a ten second breath hold, exhalation, and 45 second waiting period before repeating the process^28^. Each attack, participants performed the FUPP 4 times under continuous application guidance. The application pushed surveys at 1, 2, 24 and 48 hours.

### Outcomes

Outcomes were based on International Headache Society (IHS) guidelines^29^. Primary outcome was pain relief at 2 hours post-vaporization^29^ based on the limited supporting evidence at study initiation^18^. Secondary outcomes were 2-hour pain freedom and 2-hour most bothersome symptom (MBS) freedom^29^. Other outcomes included freedom from pain, photophobia, phonophobia, nausea, and vomiting, and rescue medication use, with data collected at 1, 2, 24 and 48 hours^29^. Pain at time 0 and each subsequent timepoint was reported as none, mild, moderate or severe^29^. Pain relief was defined as reduction of pain from moderate or severe to mild or none^29^. Pain freedom was defined as absence of pain. Sustained pain freedom was defined as pain freedom at 2 hours without return of pain or rescue medication use^29^. MBS (selected from photophobia, phonophobia, or nausea) was identified by the participant during each attack prior to vaporization; MBS freedom was defined as absence of the MBS at each subsequent timepoint. Sustained MBS freedom was defined as MBS freedom at 2 hours without return of the MBS or rescue medication use at each subsequent timepoint ^29^. All outcomes presented here were prospectively registered (NCT04360044).

### Analyses

The intention-to-treat (ITT) analysis analyzed all treated migraine attacks, counting treated attacks without recorded 2-hour data as treatment failures as per IHS guidelines^29^. The modified intention-to-treat analysis (mITT) only analyzed attacks with recorded 2-hour data. During the trial, some subjects filled out surveys early or late. We performed a sensitivity analysis analyzing only attacks with 2-hour data time stamped 1.5-3 hours post-vaporization, a time window selected to retain as much data as possible without increasing possible exposure to retention bias^30–32^. As per IHS guidelines, for all analyses, attacks for which the participant used rescue medication before the 2-hour assessment were counted as treatment failures^29^. The differences in pain relief, pain freedom, and MBS freedom among the four treatments were assessed using a generalized linear mixed effects model with a logit link for binary outcomes. A random intercept structure was included to account for the cluster effect of subjects going through the same trial multiple times. Secondary and other analyses at 1, 24, and 48 hours were conducted using similar methods. Statistical analyses were performed using RStudio, version 4.1.2 (R Foundation). All tests were two-sided with p<0.05 indicating statistical significance.

## Results

678 people were screened for eligibility, of whom 92 were enrolled (Figure 1). Participants had a median age of 41, and 82.6% were female (Table 1).

**Table 1:**
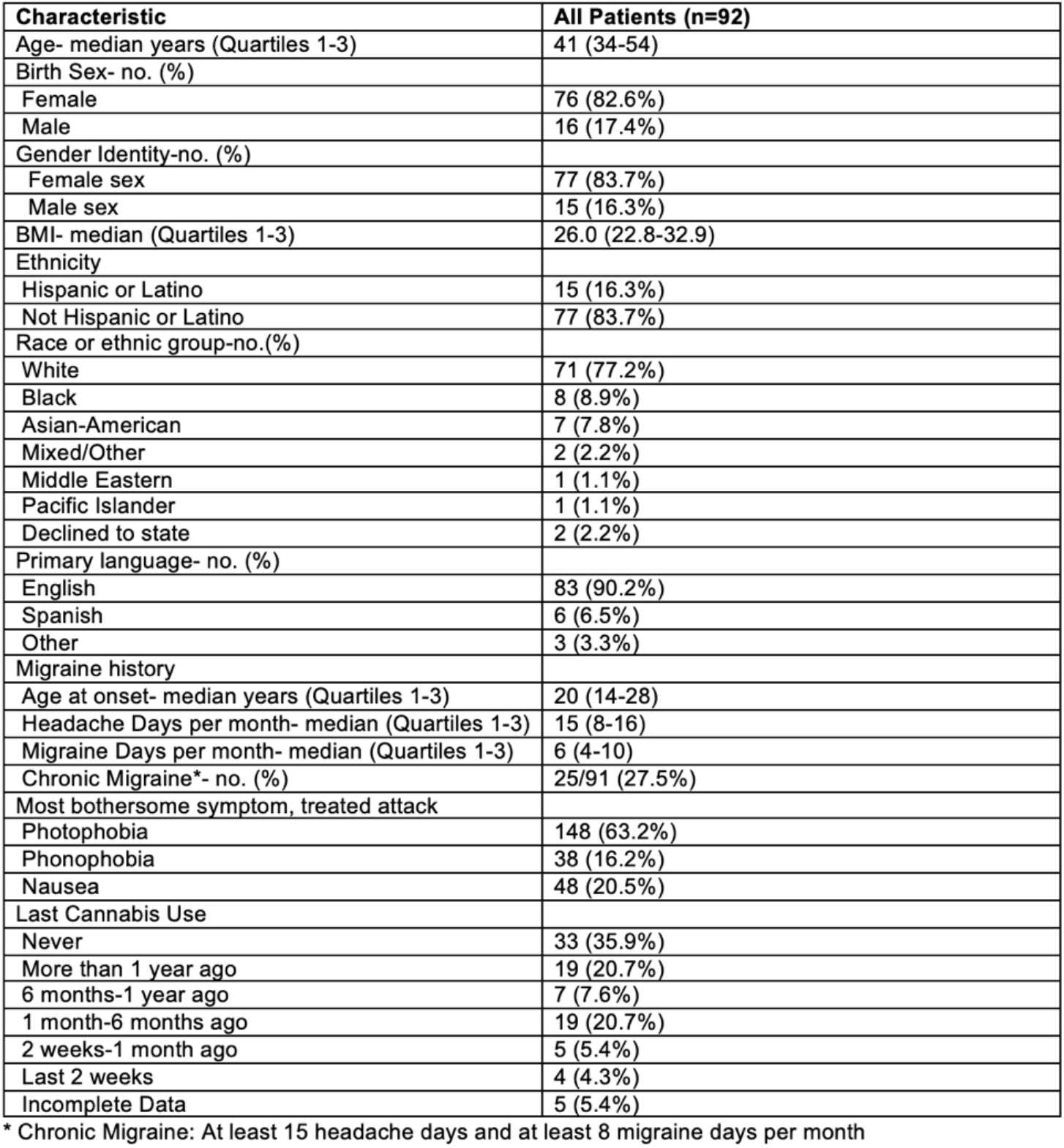
Characteristics of Patient Population.

**Figure 1:**
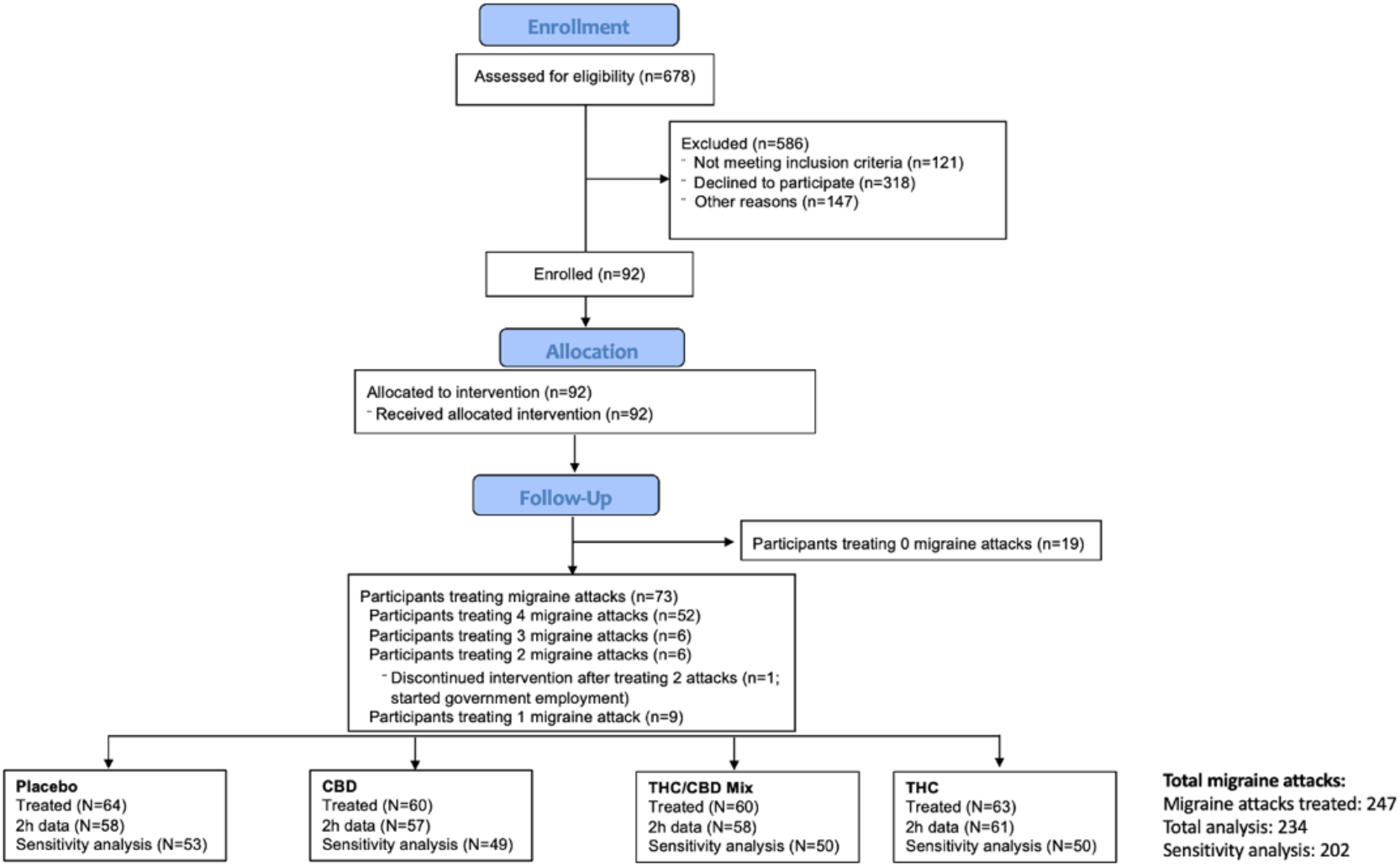
Enrollment, Randomization and Follow-up.

The ITT analysis included all 247 migraine attacks treated with vaporized cannabis from 73 participants. The mITT analysis included 234 attacks from 71 patients. The sensitivity analysis included 202 attacks from 70 patients (Figure 1). The results were similar across the ITT, mITT, and sensitivity analyses for the primary endpoint of 2-hour pain relief and secondary endpoints of 2-hour pain freedom and 2-hour MBS freedom. Where not otherwise specified, mITT results are reported as they neither used imputed data (as in the ITT) nor excluded data (as in the sensitivity analysis).

For the primary outcome of 2-hour pain relief, THC+CBD and THC-dominant were superior to placebo but CBD-dominant was not in all 3 analyses. In the ITT analysis, 2-hour pain relief responder rates were 63.9% with THC+CBD, 67.7% with THC-dominant, 50% with CBD-dominant, and 42.2% with placebo. In the mITT analysis, 2-hour pain relief was achieved by 67.2% with THC+CBD (OR [95%] 2.85 [1.22-6.65], p=0.016), 68.9% with THC-dominant (3.140 [1.35-7.30], p=0.008), 52.6% with CBD-dominant (p>0.05), and 46.6% with placebo (Figures 2 and 3). In the sensitivity analysis; 2-hour pain relief responder rates were 64.0% with THC+CBD, 70.0% with THC-dominant, 53.1% with CBD-dominant, and 45.3% with placebo.

**Figure 2:**
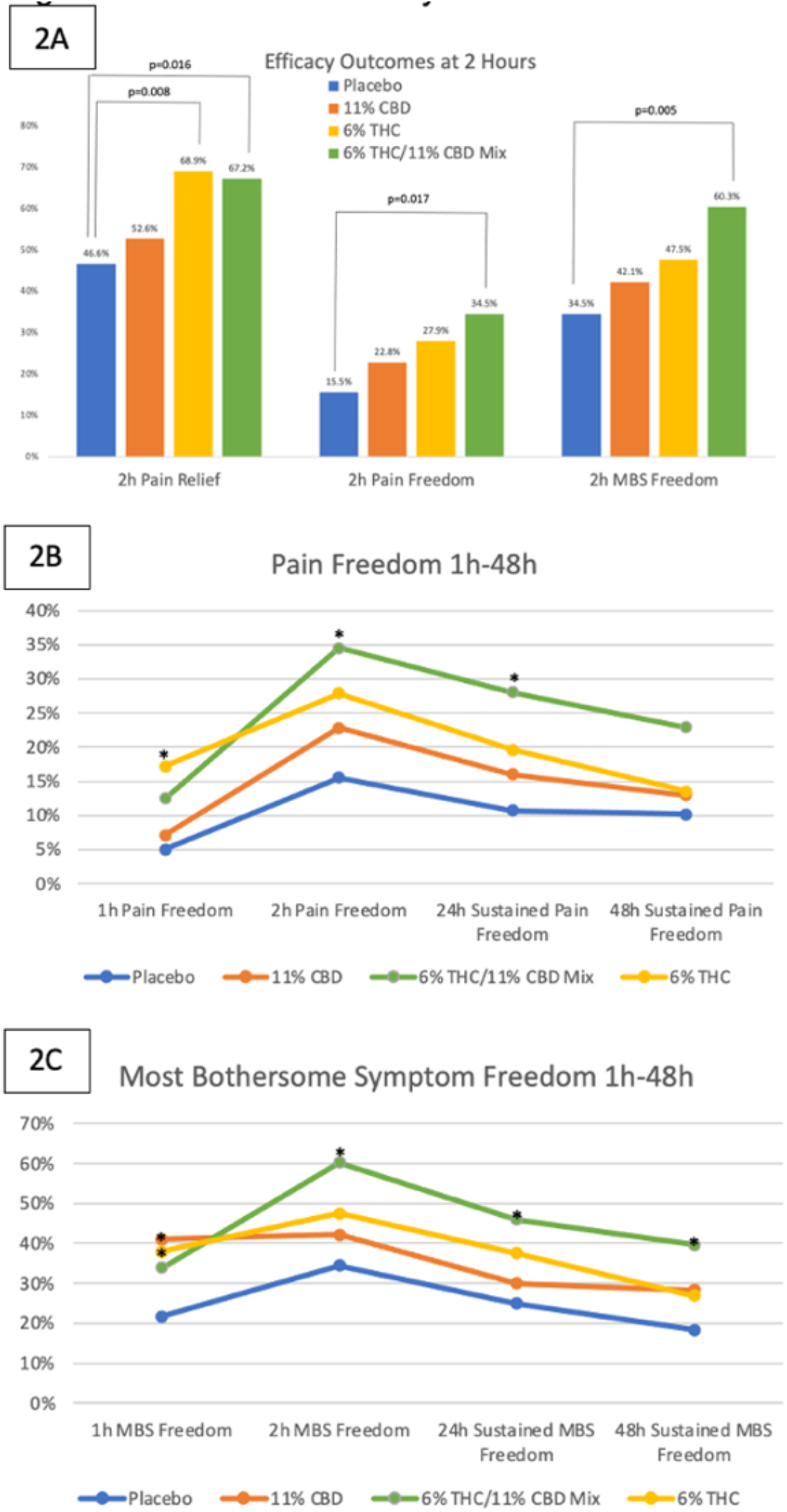
Box Plots of Efficacy Data. 2A) Efficacy Outcomes at 2 Hours. P-values were based on generalized linear mixed effects model. 2B) Pain Freedom rates at 1hand 2h andSustained Pain Freedom rates at 24h and 48h 2C) Most Bothersome Symptom (MBS) Freedom rates at 1h and 2h and Sustained MBS Freedom rates at 24h and 48h (* p<0.05)

**Figure 3:**
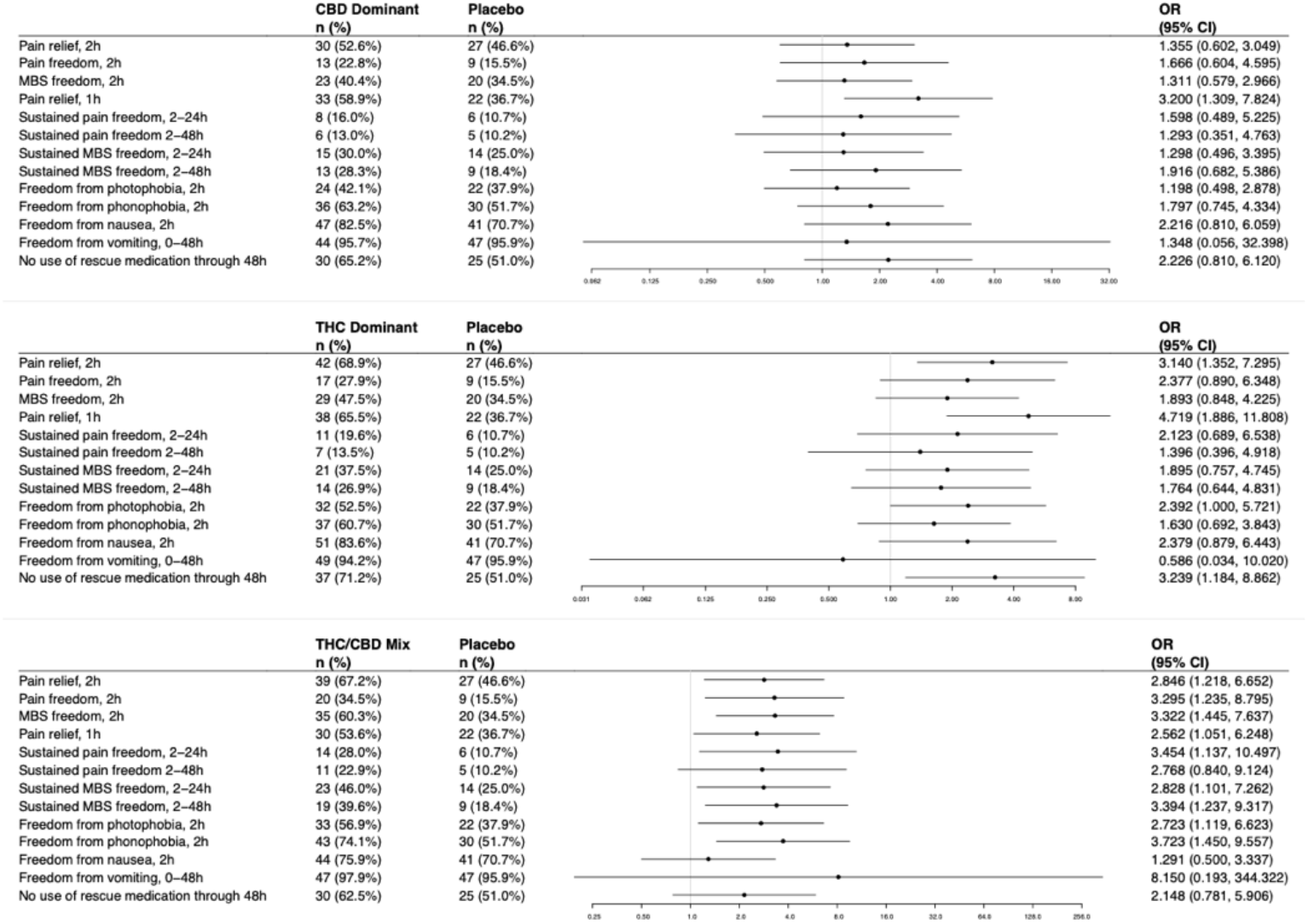
Forest Plots of Efficacy Data.

For the secondary outcomes, THC+CBD was superior to placebo in all 3 analyses for 2-hour pain freedom (mITT 34.5% vs. 15.5%, 3.30 [1.24-8.80], p=0.017) and 2-hour MBS freedom (mITT 60.3% vs. 34.5%, 3.32 [1.45-7.64], p=0.005) but THC-dominant and CBD-dominant were not (Figures 2 and 3). Adjusting for treatment session number did not affect the results of the primary or secondary outcomes.

THC+CBD was superior to placebo at 2 hours with regards to freedom from photophobia and phonophobia but not nausea or vomiting, while THC-dominant and CBD-dominant were not superior to placebo for any of these (Figure 3).

At 1 hour, pain relief responder rate for THC+CBD (53.6%, 2.56 [1.05-6.25], p=0.039), THC-dominant (65.5%, 4.72 [1.89-11.81], p<0.001), and CBD-dominant (58.9%, 3.20 [1.31-7.82], p=0.011) were all superior to 36.7% responder rate with placebo. 1-hour pain freedom was superior only for THC-dominant vs. placebo (17.2% vs 5.0%, 4.90 [1.12-21.34], p=0.034), and 1-hour MBS freedom was superior for THC-dominant (37.9%, 2.68 [1.06-6.79], p=0.038) and CBD-dominant (41.1%, 3.10 [1.21-7.91], p=0.018) but not for THC+CBD (33.9% vs., 2.19 [0.85-5.64], p=0.103) vs. placebo (21.7%).

Only THC+CBD had superior sustained benefit versus placebo (Figures 2 and 3). THC+CBD was superior to placebo regarding 24-hour sustained pain freedom (28.0% vs. 10.7%, 3.45 [1.14-10.50], p=0.029), 24-hour sustained MBS freedom (46.0% vs. 25.0%, 2.83 [1.10-7.26], p=0.031), and 48-hour sustained MBS freedom (39.6% vs. 18.4%, 3.39 [1.24-9.32], p=0.018) but the difference for 48-hour sustained pain freedom was non-significant (22.9% vs 10.2%, 2.77 [0.84-9.12], p=0.094). THC-dominant and CBD-dominant were not different from placebo regarding 24- or 48-hour sustained pain freedom or sustained MBS freedom.

Adverse events are reported in Table 2. Mean subjective highness on a 0-10 scale at 1 hour was greatest with THC-dominant at 3.5 (SD 3.0), followed by 2.4 (SD 2.5) with THC+CBD, 1.5 (SD 2.0) with CBD-dominant, and 0.6 (SD 1.20) with placebo. At 2 hours subjective highness was reduced to 2.4 (SD 2.8) with THC-dominant, 1.3 (SD 1.9) with THC+CBD, 0.9 (SD 1.6) with CBD-dominant, and 0.4 (SD 1.0) with placebo. THC-dominant had the greatest euphoria, cognitive impairment and subjective highness, followed by THC+CBD, then CBD-dominant, and least with placebo. Across all 4 treatments there were no serious adverse events (Table 2).

**Table 2:** Adverse Events. Table 2A) Rates of queried adverse events Table 2B) Categorized free response adverse events

| 2A | Placebo | CBD | THC+CBD | THC |
| --- | --- | --- | --- | --- |
| **1 Hour (N)** | N=60 | N=56 | N=56 | N=58 |
| Sleepiness | 16 (26.7%) | 21 (37.5%) | 25 (44.6%) | 24 (41.4%) |
| Euphoria | 4 (7%) | 5 (8.9%) | 16 (28.6%) | 21 (36.2%) |
| Cognitive Impairment | 4 (7%) | 8 (14.3%) | 12 (21.4%) | 20 (34.5%) |
| Highness (0-10) Mean (SD) | 0.6 (1.2) | 1.5 (2.0) | 2.4 (2.5) | 3.5 (3.0) |
| **2 Hours (N)** | N=58 | N=57 | N=58 | N=61 |
| Sleepiness | 22 (37.9%) | 29 (50.9%) | 18 (31.0%) | 26 (42.6%) |
| Euphoria | 1 (1.7%) | 2 (3.5%) | 6 (10.3%) | 17 (27.9%) |
| Cognitive Impairment | 3 (5.2%) | 4 (7.0%) | 7 (12.1%) | 17 (26.2%) |
| Highness (0-10) Mean (SD) | 0.4 (1.0) | 0.9 (1.6) | 1.3 (1.9) | 2.4 (2.8) |

| 2B | Placebo | CBD | THC+CBD | THC |
| --- | --- | --- | --- | --- |
| **Participants reporting any adverse events** | **3/60 (5.0%)** | **11/56 (19.6%)** | **11/56 (19.6%)** | **18/58 (31.0%)** |
| **Adverse events reported at 1 hour** | **0** | **7** | **12** | **13** |
| Sedation | 0 | 2 | 3 | 0 |
| Slowness | 0 | 1 | 0 | 4 |
| Paresthesias | 0 | 0 | 1 | 2 |
| Throat irritation | 0 | 3 | 2 | 0 |
| Increased appetite | 0 | 2 | 1 | 1 |
| Impaired cognition | 0 | 0 | 2 | 1 |
| Very high | 0 | 0 | 0 | 1 |
| Impaired focus | 0 | 0 | 2 | 2 |
| Dizziness | 0 | 0 | 0 | 2 |
| Dry mouth | 0 | 2 | 1 | 0 |
| **Adverse events reported by ≥1% of participants at 2 hours** | **3/58 (5.2%)** | **4/57 (7.0%)** | **4/58 (6.9%)** | **11/61 (18.0%)** |
| Sedation | 0 | 3 | 2 | 2 |
| Impaired focus | 1 | 0 | 0 | 1 |
| Slowness | 0 | 0 | 0 | 3 |
| Dizziness | 0 | 0 | 2 | 2 |
| Throat irritation | 0 | 1 | 1 | 0 |
| Dry mouth | 2 | 0 | 1 | 0 |
| Paresthesias | 0 | 0 | 0 | 1 |
| Increased appetite | 0 | 0 | 0 | 1 |
| Impaired cognition | 0 | 0 | 1 | 1 |
| Very high | 0 | 0 | 0 | 0 |

Regarding blinding, correct identification of treatment at 2 hours was 29.3% (17/58) with placebo, 10.5% (6/57) with CBD-dominant, 13.1% (8/61) with THC-dominant, and 20.7% (12/58) with THC+CBD. Participants selected “unknown” 24% of the time; 28% of the time with placebo, 26% with CBD-dominant, 22% with THC+ CBD, and 21% with THC-dominant. While 50% of treatments could have contained THC (THC-dominant or THC+CBD) and in this study 50.9% (119/234) treatments contained THC, participants correctly identified that a treatment contained THC (selecting “THC-dominant”, “THC+CBD”, or “Either THC-dominant or THC+CBD”) slightly less than expected by chance, 46.6% (27/58) of the time after THC+CBD treatment and 49.2% (30/61) after THC-dominant treatment. Meanwhile, 32% of participants selected a THC-containing answer after treatment with CBD-dominant and 14% after treatment with placebo.

## Discussion

In this study, the first randomized, double-blind, placebo-controlled trial testing the efficacy of cannabinoids for acute migraine, 6% THC+11% CBD was superior to placebo for pain relief, pain freedom and MBS at 2 hours, as well as freedom from photophobia and phonophobia at 2 hours and 24-hour sustained pain freedom and sustained MBS freedom and 48-hour sustained MBS freedom.

THC potencies (6%) were lower than typical with cannabis available via American dispensaries and subjective highness (2.4 out of 10 at 1 hour) with THC+CBD was roughly half of that reported by research participants using cannabis ad lib^33^, bolstering evidence that higher potencies and titrating to highness are unnecessary for medicinal benefit. THC+CBD had less euphoria, subjective cognitive impairment, and subjective highness than THC-dominant. CBD is reported to be a non-competitive, negative allosteric modulator of the CB1 receptor, by which CBD may decrease the psychoactive effects of THC^34–36^.

The blinding data, as well as THC+CBD having more acute migraine benefit with less psychoactive effects as compared to THC-dominant, suggests that this study’s findings are not explainable by expectation effects or unmasking due to psychoactive side effects.

THC+CBD’s efficacy for reducing photophobia and phonophobia but not nausea in this study may be in part explainable by lower time 0 rates of nausea (59.9%) than photophobia (91.9%) and phonophobia (87.0%) in this study. That THC+CBD reduced photophobia and phonophobia but not nausea or vomiting in this study demonstrates that THC+CBD’s effects on MBS are not explainable by THC’s established antiemetic effects^37^ (Figure 3).

Strengths of the study include generalizability by enrolling patients with both episodic (72.5%) and chronic (27.5%) migraine to best reflect the larger patient population; historically most acute migraine studies have excluded patients with chronic migraine. Limitations of this study include that the study only examined THC and CBD; minor cannabinoids and terpenes were not studied. It also only studied single potencies of THC of CBD and a single THC:CBD ratio.

Typical of acute migraine RCTs, this study did not assess repeated administrations or regular, long-term treatments to assess possible risks (including rates of development of medication overuse headache and cannabis use disorder) or benefits (including possible migraine preventive effects)^29,38^. A small body of literature shows improvements on patient reported outcomes when using cannabis-based medicinal products on a regular, longer term or preventive basis^20,39^. More research is needed to evaluate repeated administrations and regular, long-term use of cannabinoids for migraine.

## Conclusion

In this first randomized, double-blind, placebo-controlled trial testing the efficacy of cannabinoids for the acute treatment of migraine, vaporized 6% THC+11% CBD cannabis flower was superior to placebo for pain relief, pain freedom, and MBS freedom at 2 hours as well as 24-hour sustained pain freedom and sustained MBS freedom and 48-hour sustained MBS freedom. Future research should include multicenter studies and long-term studies of benefits and risks with repeated use.

## Data Availability

All data produced in the present study are available upon reasonable request to the authors

## Acknowledgements

This study was sponsored by the Migraine Research Foundation, now administered by Hartford HealthCare. The project described was partially supported by the UCSD Academic Senate and the National Institutes of Health, Grant UL1TR001442. The content is solely the responsibility of the authors and does not necessarily represent the official views of the NIH. Study drug was provided by the National Institute on Drug Abuse Drug Supply Program. The authors thank the National Institute on Drug Abuse Drug Supply Program and the National Center for Natural Products Research at the University of Mississippi School of Pharmacy. Vaporizers were donated by Storz & Bickel GmbH & Co. KG, which did not provide any financial support of this study and was not involved in the design or conduct of the study or manuscript preparation. Thanks to Dr. Brooke Towne, MD, Phirum Nguyen, Gayle Dizon, Jay Patel, Brett Taylor, and Megan Sweeney for assistance with conducting this study. Thank you to all the participants who volunteered for this study.

